# Health related quality of life of oral cancer patients who receive radiotherapy with or without chemotherapy in a tertiary referral centre in Sri Lanka- A prospective study

**DOI:** 10.1101/2022.07.27.22278132

**Authors:** Shamini Kosgallana, Prasanna Jayasekara, Prasad Abeysinghe, Ratilal Lalloo

## Abstract

**Purpose:** To assess the health-related quality of life (HRQOL) of oral cancer patients who receive radiotherapy (RT) with or without chemotherapy and the changes in HRQOL.

**Patients and Methods:** A prospective study was conducted among oral cancer patients who receive RT with or without chemotherapy. Two European Organization for the Research and Treatment of Cancer Quality of Life Questionnaires: EORTC QLQ-C30 and EORTC QLQ-H&N35, were used to assess HRQOL before RT, during the last week of RT and three months after RT. High scores of symptom domains and items indicate low HRQOL and wise versa for functional and ‘*Global health status’* domains.

**Results:** Ninety oral cancer patients were included. The majority of the sample were males (88%) and 68% were aged 50-69. The median scores of all the domains and items in EORTC QLQ-H&N35 and symptom domains and items in EORTC QLQ-C30 were higher during the last week of RT compared to the baseline. The functional domains of EORTC QLQ-C30 showed the highest median values (100.0) at baseline but much less values (<66.7) during last week of RT. Except for ‘*Appetite loss’* item, all the symptom domains and items scores were lower at three months after RT than the median scores during last week of RT. Statistically significant differences were observed in almost all the changes in HRQOL between three-time frames (p<0.05).

**Conclusions:** HRQOL of oral cancer patients declined due to RT from baseline to the last week of RT and improved three months after RT from last week of RT but had not returned to the baseline level.

## Introduction

The majority of the head and neck cancers are oral cancers.^1,2,3^ Oral cancer as defined by the American Joint Committee on Cancer and the Union for International Cancer Control in the tumor-node-metastasis staging classification, includes carcinomas of the oral cavity originating from the mucosal lip, anterior two-thirds of the tongue, buccal mucosa, floor of mouth, hard palate, lower and upper alveolus and gingiva, and the retromolar trigone.^4^ Oral cancers account for the third highest standardized death rate in countries with low and medium human development index. Globally, Papua New Guinea had the highest age-specific incidence rate for oral cancer with 21.2 and 12 cases per 100,000 population for males and females respectively.^5^ Oral cancer is ranked the number one cancer among males in Sri Lanka, with an age-standardize incidence rate of 19.1 per 100,000 people in 2019. As elsewhere, in Sri Lanka, it is more common in men than in women, comprising 15% and 3% of all cancers in males and females respectively.^6^

There are many different modalities available for treating oral cancers. Surgery, radiotherapy (RT) and chemotherapy alone or in combination are recommended for treating oral cancers.^7,8^ RT with or without chemotherapy is used as the primary treatment modality in early stage and un-resectable tumours to avoid anticipated functional and cosmetic defects. RT is also used in patients when the surgery can be high risk due to comorbidities or poor performance capacity to withstand a surgery, in recurrent malignancies and when patient’s preference is RT.^9^ In Sri Lanka, RT is commonly used in oral cancer patients post surgically. As most of the oral cancer patients are between 50 and 70 years of age, RT is also used alone or with chemotherapy.^10^

Although treatments for cancer are employed to improve the patient’s health related quality of life (HRQOL), they are associated with several side effects which deteriorate patients’ HRQOL.^11^ Dermatitis, dysphagia, mucositis, loss of taste, xerostomia, osteoradionecrosis, trismus, candidiasis and radiation caries are some of the commonest side effects.^8,12,13^ There are generic and disease specific tools to assess HRQOL. Disease-specific measures have greater sensitivity than generic measures because they were developed for specific conditions to measure symptoms and impacts associated with that condition.^14^ The European Organization for the Research and Treatment of Cancer (EORTC) Quality of Life Questionnaire-C30 (EORTC QLQ-C30), assesses details on five functional domains, one global health status domain, three symptom domains and six items. This allows detailed information about how the various domains are influenced by the disease and treatment and overall quality-of -life.^10^ The EORTC quality-of-life group has developed a site specific tool to measure HRQOL for head and neck cancers (EORTC QLQ-H&N35). The combination of these two questionnaires provide information on the perception of the patient, the impact of the disease and its treatment, side effects of the treatments, impact of medical interventions, as well as its performance concerning various aspects of life.^15^

It has been emphasized that the need of prospective studies to obtain insight into the relation between pretreatment HRQOL and outcome after treatment, and the relationship between changes in HRQOL and functioning over the period of time.^16^ Therefore, it is important to assess the changes of HRQOL due to their current treatment modalities. The aim of this research was to assess the HRQOL in oral cancer patients before RT with or without chemotherapy, during the last week of the RT course and three months following the completion of RT, and the changes in HRQOL during these time periods.

## Methods

A prospective longitudinal study was carried out among oral cancer patients. The patients were recruited from outpatient clinics and hospitalized patients at the Apeksha hospital, Maharagama, Colombo, Sri Lanka. Biopsy confirmed adult oral cancer patients who were newly diagnosed during the last three months and were waiting for RT with or without chemotherapy as the initial treatment, and at any stage were included in this study. Patients who had already undergone surgical treatment and patients who receive small doses of radiation therapy as a palliative care were excluded from the study.

After confirming the eligibility criteria, the purpose and the objectives of the study were explained to the patients. The informed written consent was obtained before collecting the data. The short interviewer-administered questionnaire was used to gather socio-demographic data. The clinical information of the patients was obtained from clinical records.

Validated self-administered EORTC QLQ-C30 and EORTC QLQ-H&N35 were used to gather patient-reported outcomes regarding HRQOL at baseline.^17^ The patients were followed up and the same two questionnaires were completed again by the same patients during last week of RT course and three months after completion of RT. Ethical approval to conduct the study was obtained from the Ethics Review Committee of Faculty of Medicine, University of Colombo, Sri Lanka (EC-15-200).

### HRQOL questionnaires

HRQOL was assessed by EORTC QLQ-H&N35 consisting of seven symptom domains and 11 single symptom items, and EORTC QLQ-C30 consisted of one ‘*Global health status*’ domain, five functional domains, three symptom domains and six single symptom items. These two questionnaires were validated previously and used in cancer patients in Sri Lanka.^17^ All the responses in the questionnaires had a four-point Likert scale, namely “Not at all”, “A little”, “Quite a bit” and “Very much” except for the two items for the ‘*Global health status*’ domain, which has seven Likert scales ranging from “very poor” to “excellent” in the EORTC QLQ-C30 questionnaire.

### Data analysis

Data entry and analysis were carried out using version 22 of the Statistical Package of Social Sciences. All of the domains and single-item measures range in score from zero to 100. A high score for a functional domain represents a high or healthy level of functioning and a high score for the ‘*Global health status’* represents a high HRQOL. A high score for symptom domains and items represents a high level of symptoms or problems.

Descriptive analysis was performed to present socio-demographic data and clinical characteristics of the study sample. Normality of the data was assessed by observing histograms, skewness and using Kolmogorov-Smirnov and Shapiro-Wilk statistical tests. The distributions of data were not normal, and all the analyses were performed using non-parametric tests. Median scores were used for analysis, and inter-quartile range (IQR), the mean scores and standard deviation (SD) were presented in the results for better understanding and for comparison with other studies.

The change in HRQOL due to RT was calculated by deducting baseline scores from the scores during last week of RT, baseline scores from the scores three months after RT, the scores during last week of RT from the scores of three months after RT.

The significance of the changes in HRQOL was tested using Wilcoxon Signed Ranks Test and a p-value <0.05 was considered statistically significant.

## Results

At baseline the sample size was 90, during the last week of RT 86 patients, and at three months after RT 75 patients were followed-up. The majority of the sample (68%) was 50 to 69 years of age. Nearly 88% of the sample consisted of males and 91% were married. When considering the site of the oral cancer, 40% had cancer in the anterior 2/3 of the tongue and 22% on the buccal mucosa. Of the sample, 72% were in the late stage of the disease at diagnosis and 63% were treated with chemo and RT. Only 19% of the sample had diseases other than oral cancer (Table 1).

**Table 1:** Distribution of the Study Population by Socio Demographic and Clinical Characteristics (n=90)

| Characteristics | Frequency | Percentage (%) |
| --- | --- | --- |
| **Age** |  |  |
| 35-49 | 14 | 15.5 |
| 50-69 | 61 | 67.8 |
| >70 | 15 | 16.7 |
| **Sex** |  |  |
| Female | 11 | 12.2 |
| Male | 79 | 87.8 |
| **Civil status** |  |  |
| Married | 82 | 91.1 |
| Unmarried | 8 | 8.9 |
| **Education level** |  |  |
| Up to grade 5 | 28 | 31.1 |
| Up to O/L | 55 | 61.1 |
| Up to A/L | 6 | 6.7 |
| Diploma/ Degree | 1 | 1.1 |
| **Employment status** |  |  |
| Unemployed | 18 | 20.0 |
| Self employed | 55 | 61.1 |
| Employed | 9 | 10.0 |
| Pensioner | 8 | 8.9 |
| **Income** |  |  |
| <15000 | 37 | 41.1 |
| 15000-30000 | 42 | 46.7 |
| > 30000 | 11 | 12.2 |
| **Site of the oral cancer** |  |  |
| Lip | 2 | 2.2 |
| Anterior two-thirds of the tongue | 36 | 40.0 |
| Buccal mucosa | 20 | 22.2 |
| Floor of the mouth | 12 | 13.3 |
| Hard palate | 5 | 5.6 |
| Lower and upper alveolar ridge | 1 | 1.1 |
| Retromolar trigone | 8 | 8.9 |
| More than two sites* | 6 | 6.7 |
| **Stage** |  |  |
| Early stage (stage I and II) | 23 | 25.6 |
| Late stage (stage III and IV) | 65 | 72.2 |
| Missing | 2 | 2.2 |
| **Metastasis** |  |  |
| None | 37 | 41.1 |
| Lymph node | 49 | 54.4 |
| Systemic | 2 | 2.2 |
| Missing | 2 | 2.2 |
| **Treatment modality** |  |  |
| Radiotherapy | 33 | 36.7 |
| Chemo-radiotherapy | 57 | 63.3 |
| **Other diseases** |  |  |
| None | 73 | 81.1 |
| Any other disease ** | 17 | 18.9 |

\* Anterior two-thirds of the tongue and floor of mouth=4, Hard palate, buccal mucosa and alveolus=1, Buccal mucosa and floor of the mouth=1
\*\*Diabetes=4, Hypertension=4, Diabetes and hypertension=2, Asthma=2, Arthritis= 2, Gastritis=2, Cerebrovascular diseases=1

Most of the domain and item median scores of EORTC QLQ-H&N35 were zero at baseline. During the last week of RT all the median scores were more than 50.0 except ‘*Teeth’* (33.3, IQR 0.0-66.7), *‘Coughing’* (33.3, IQR 0.0-33.3*), ‘Feeding tube’* (0.0. IQR 0.0-100.0) and ‘*Weight gain’* (0.0, IQR 0.0-0.0) items. Furthermore, mean scores for all the parameters of EORTC QLQ-H&N35 were higher during the last week of RT compared to baseline. The most pronounced symptom items were *‘Dry mouth’* (89.9, SD 2.2), *‘Sticky saliva’* (89.9, SD 2.4), *‘Pain killers’* (97.7, SD 1.6), *‘Nutritional supplements’* (98.8, SD 1.2) and *‘weight loss’* (94.2, SD 2.5) during last week of RT. Median scores for *‘Speech problem’*, ‘*Less sexuality’* and ‘*Opening mouth*’ remained the same from baseline to three months after RT, while other scores were higher. Almost all the domain and item median scores were lower at three months after RT than the last week of RT except for *‘Pain killers’* and ‘*Nutritional supplements’*, which remained the same (Table 2).

**Table 2:** Comparison of EORTC QLQ-H&N35 Scores of the Sample at Baseline, During Last Week of RT Course and three Months After RT.

| Domains and items | Baseline(n=90) |  | Last week of RT(n=86) |  | Three months after RT(n=75) |  |
| --- | --- | --- | --- | --- | --- | --- |
|  | Mean (SD) | Median (IQR) | Mean (SD) | Median (IQR) | Mean (SD) | Median (IQR) |
| Pain | 14.6<br>(1.6) | 8.3<br>(0.0-25.0) | 66.2<br>(2.9) | 66.7<br>(47.9-91.7) | 22.7<br>(2.8) | 16.7<br>(0.0-33.3) |
| Swallowing | 9.7<br>(1.5) | 4.2<br>(0.0-16.7) | 56.8<br>(2.0) | 58.3<br>(50.0-68.8) | 27.7<br>(2.9) | 16.7<br>(8.3-50.0) |
| Senses problem | 8.5<br>(1.2) | 0.0<br>(0.0-16.7) | 52.1<br>(1.2) | 50.0<br>(50.0-50.0) | 35.3<br>(1.8) | 33.3<br>(33.3-50.0) |
| Speech problem | 6.9<br>(1.6) | 0.0<br>(0.0-11.1) | 54.0<br>(2.8) | 55.6<br>(44.4-66.7) | 20.4<br>(3.3) | 0.0<br>(0.0-44.4) |
| Trouble with social eating | 13.0<br>(1.9) | 8.3<br>(0.0-16.7) | 72.8<br>(2.1) | 75.0<br>(58.3-91.7) | 43.4<br>(3.4) | 33.3<br>(16.7-66.7) |
| Trouble with social contact | 4.3<br>(1.3) | 0.0<br>(0.0-0.0) | 58.5<br>(2.8) | 56.7<br>(40.0-80.0) | 27.1<br>(3.4) | 20.0<br>(0.0-53.3) |
| Less sexuality | 3.9<br>(1.9) | 0.0<br>(0.0-0.0) | 46.2<br>(6.61) | 66.7<br>(0.0-100.0) | 16.7<br>(5.4) | 0.0<br>(0.0-33.3) |
| Teeth | 13.0<br>(2.0) | 0.0<br>(0.0-33.3) | 34.5<br>(3.3) | 33.3<br>(0.0-66.7) | 40.2<br>(3.9) | 33.3<br>(0.0-66.7) |
| Opening mouth | 20.4<br>(3.1) | 0.0<br>(0.0-33.3) | 59.7<br>(3.5) | 66.7<br>(33.3-100.0) | 29.3<br>(4.2) | 0.0<br>(0.0-66.7) |
| Dry mouth | 15.2<br>(2.4) | 0.0<br>(0.0-33.3) | 89.0<br>(2.2) | 100.0<br>(66.7-100.0) | 75.6<br>(3.8) | 100.0<br>(66.7-100.0) |
| Sticky saliva | 16.7<br>(2.3) | 0.0<br>(0.0-33.3) | 89.8<br>(2.4) | 100.0<br>(100.0-100.0) | 62.2<br>(4.3) | 66.7<br>(33.3-100.0) |
| Coughing | 4.4<br>(1.2) | 0.0<br>(0.0-0.0) | 26.7<br>(2.5) | 33.3<br>(0.0-33.3) | 8.9<br>(2.1) | 0.0<br>(0.0-0.0) |
| Felt ill | 8.6<br>(2.0) | 0.0<br>(0.0-0.0) | 66.7<br>(3.2) | 66.7<br>(66.7-100.0) | 36.9<br>(4.1) | 33.3<br>(0.0-66.7) |
| Pain killers | 35.6<br>(5.1) | 0.0<br>(0.0-100.0) | 97.7<br>(1.6) | 100.0<br>(100.0-100.0) | 53.3<br>(5.8) | 100.0<br>(0.0-100.0) |
| Nutritional supplements | 18.9<br>(4.2) | 0.0<br>(0.0-0.0) | 98.8<br>(1.2) | 100.0<br>(100.0-100.0) | 77.3<br>(4.9) | 100.0<br>(100.0-100.0) |
| Feeding tube | 1.1<br>(1.1) | 0.0<br>(0.0-0.0) | 39.5<br>(5.3) | 0.0<br>(0.0-100.0) | 22.7<br>(4.9) | 0.0<br>(0.0-0.0) |
| Weight loss | 23.3<br>(4.5) | 0.0<br>(0.0-0.0) | 94.2<br>(2.5) | 100.0<br>(100.0-100.0) | 30.7<br>(5.4) | 0.0<br>(0.0-100.0) |
| Weight gain | 0.0<br>(0.0) | 0.0<br>(0.0-0.0) | 4.7<br>(2.8) | 0.0<br>(0.0-0.0) | 1.3<br>(1.3) | 0.0<br>(0.0-0.0) |

For EORTC QLQ-C30, the median scores for all functional domains were 100.0 and zero for all the symptom domains and single items at baseline. The median score for the *‘Global health status’* domain was 83.3 (IQR 83.3-100.0) at baseline. All the symptom domains and item median values had increased from baseline to the last week of RT, except ‘*Nausea and vomiting’* and *‘Appetite loss’*, which remained the same. ‘*Physical’*, ‘*Role’*, ‘*Cognitive’* and *‘Social’ functioning* showed more impairment at three months after RT compared to the baseline. Except for the ‘*Appetite loss’* item score, all the symptom domain and item scores were lower at three months after RT compared to the last week of RT (Table 3).

**Table 3:**
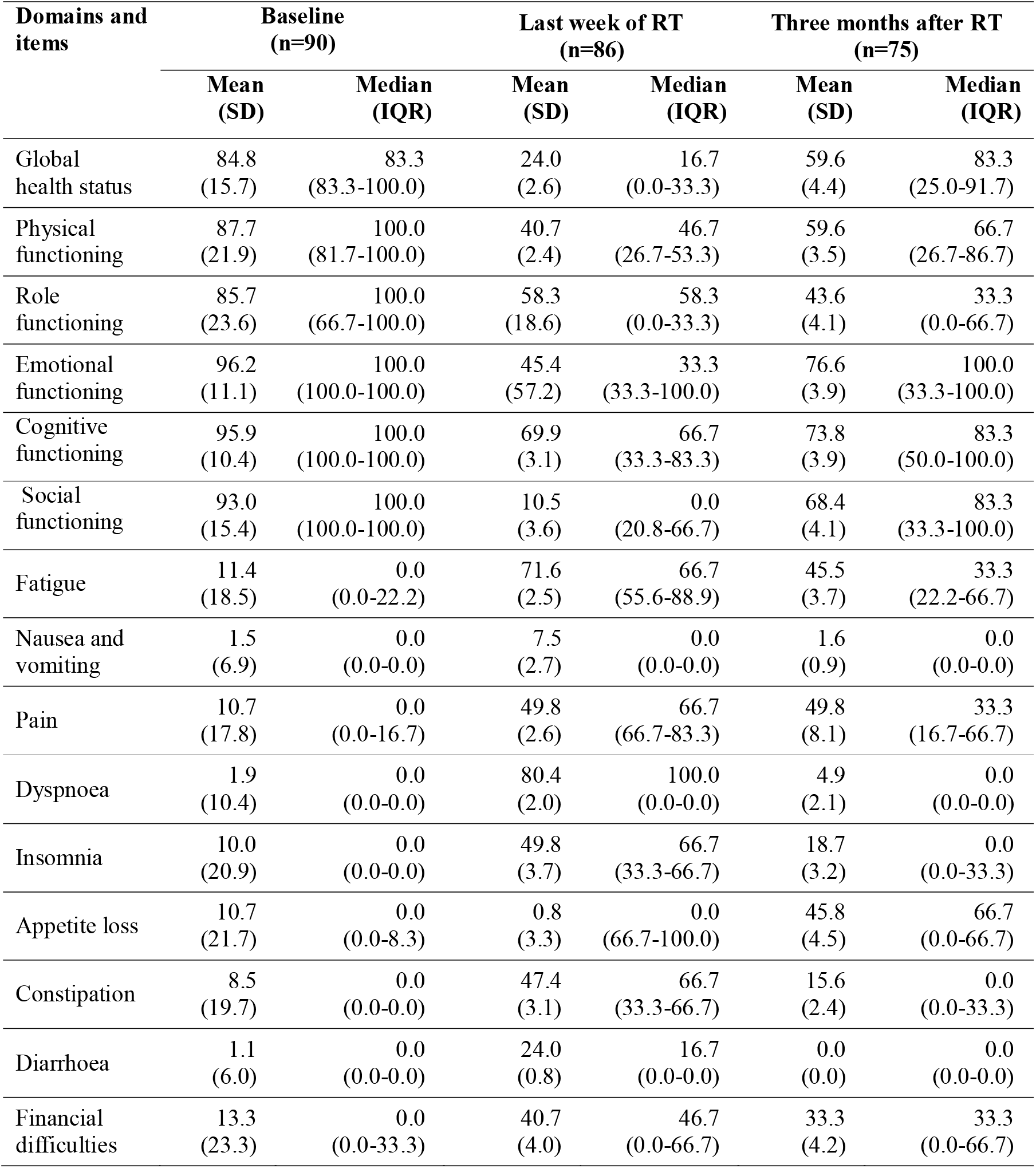
Comparison of EORTC QLQ-C30 Scores of the Sample at Baseline, During Last Week of RT Course and Three Months After RT.

The changes in HRQOL assessed by EORTC QLQ-H&N35 and their significance are tabulated in Table 4. The change of median scores for all the domains and items were significant from baseline to last week of RT, from baseline to three months after RT and from last week of RT to three months after RT (p<0.05) except for the ‘*Weight gain’* for all three changes and change of *‘Weight loss’* from baseline to three months after RT.

**Table 4:** Comparison of Changes of EORTC QLQ-H&N35 Scores (Change of OHRQOL) of the Sample.

| Domains and items | Baseline to last week of RT course (n=86) |  | Baseline to three months after RT(n=75) |  | Last week of RT course to three months after RT(n=75) |  |
| --- | --- | --- | --- | --- | --- | --- |
|  | Median (IQR) | Sig.<sup>a</sup> | Median (IQR) | Sig.<sup>a</sup> | Median (IQR) | Sig.<sup>a</sup> |
| Pain | 50.0<br>(33.3-75.0) | Z=-7.9<sup>b</sup><br>**P=0.000** | 0.0<br>(-8.3-25.0) | Z= -2.1<sup>b</sup><br>**P=0.035** | -50.0<br>(-66.7--25.0) | Z=-7.1<sup>c</sup><br>**P=0.000** |
| Swallowing | 50.0<br>(33.3-58.3) | Z=-8.0<sup>b</sup><br>**P=0.000** | 8.3<br>(0.0-41.7) | Z=-5.5<sup>b</sup><br>**P=0.000** | -25.0<br>(-50.0--8.3) | Z=-6.6<sup>c</sup><br>**P=0.000** |
| Senses problem | 50.0<br>(33.3-50.0) | Z=-8.2<sup>b</sup><br>**P=0.000** | 33.3<br>(16.7-33.3) | Z=-7.0<sup>b</sup><br>**P=0.000** | -16.7<br>(-33.3-0.0) | Z=-5.9<sup>c</sup><br>**P=0.000** |
| Speech problem | 50.0<br>(33.3-66.7) | Z=-7.7<sup>b</sup><br>**P=0.000** | 0.0<br>(0.0-0.0) | Z=-4.3<sup>b</sup><br>**P=0.000** | -33.3<br>(-55.6- -11.1) | Z=-6.4<sup>c</sup><br>**P=0.000** |
| Social eating | 66.7<br>(47.9-75.0) | Z=-8.0<sup>b</sup><br>**P=0.000** | 33.3<br>(8.3-50.0) | Z=-6.6<sup>b</sup><br>**P=0.000** | -25.0<br>(-50.0--8.3) | Z=-6.2<sup>c</sup><br>**P=0.000** |
| Social contact | 53.3<br>(38.3-73.3) | Z=-7.9<sup>b</sup><br>**P=0.000** | 20.0<br>(0.0-46.7) | Z=-5.7<sup>b</sup><br>**P=0.000** | -33.3<br>(-60.0- -0.0) | Z=-5.3<sup>c</sup><br>**P=0.000** |
| Less sexuality | 33.3<br>(0.0-91.7) | Z=-4.4<sup>b</sup><br>**P=0.000** | 0.0<br>(0.0-25.0) | Z=-2.6<sup>b</sup><br>**P=0.010** | -33.3<br>(-66.7--0.0) | Z=-3.8<sup>c</sup><br>**P=0.000** |
| Teeth | 33.3<br>(0.0-33.3) | Z=-5.1<sup>b</sup><br>**P=0.000** | 33.3<br>(0.0-66.7) | Z=-5.1<sup>b</sup><br>**P=0.000** | 0.0<br>(0.0- -33.3) | Z=-2.4<sup>b</sup><br>**P=0.018** |
| Opening mouth | 33.3<br>(33.3-66.7) | Z=-7.0<sup>b</sup><br>**P=0.000** | 0.0<br>(0.0-33.3) | Z=-2.2<sup>b</sup><br>**P=0.026** | -33.3<br>(-33.3-0.0) | Z=-5.5<sup>c</sup><br>**P=0.000** |
| Dry mouth | 66.7<br>(66.7-100.0) | Z=-8.1<sup>b</sup><br>**P=0.000** | 66.7<br>(33.3-100.0) | Z=-7.1<sup>b</sup><br>**P=0.000** | 0.0<br>(-33.3-0.0) | Z=-3.1<sup>c</sup><br>**P=0.002** |
| Sticky saliva | 66.7<br>(66.7-100.0) | Z=-7.9<sup>b</sup><br>**P=0.000** | 33.3<br>(33.3-66.7) | Z=-6.5<sup>b</sup><br>**P=0.000** | -33.3<br>(-66.7-0.0) | Z=-4.8<sup>c</sup><br>**P=0.000** |
| Coughing | 33.3<br>(0.0-33.3) | Z=-5.9<sup>b</sup><br>**P=0.000** | 0.0<br>(0.0-0.0) | Z=-1.7<sup>b</sup><br>P=0.088 | -33.3<br>(-33.3-0.0) | Z=-5.6<sup>c</sup><br>**P=0.000** |
| Felt ill | 66.7<br>(33.3-66.7) | Z=-7.7<sup>b</sup><br>**P=0.000** | 33.3<br>(0.0-33.3) | Z=-5.2<sup>b</sup><br>**P=0.000** | -33.3<br>(-66.7-0.0) | Z=-5.2<sup>c</sup><br>**P=0.000** |
| Pain killers | 100.0<br>(0.0-100.0) | Z=-7.3<sup>b</sup><br>**P=0.000** | 0.0<br>(0.0-100.0) | Z=-2.7<sup>b</sup><br>**P=0.008** | 0.0<br>(-100.0-0.0) | Z=-5.8<sup>c</sup><br>**P=0.000** |
| Nutritional Suppl. | 100.0<br>(100.0-100.0) | Z=-8.3<sup>b</sup><br>**P=0.000** | 100.0<br>(0.0-100.0) | Z=-6.4<sup>b</sup><br>**P=0.000** | 0.0<br>(0.0-0.0) | Z=-4.0<sup>c</sup><br>**P=0.000** |
| Feeding tube | 0.0<br>(0.0-100.0) | Z=-5.8<sup>b</sup><br>**P=0.000** | 0.0<br>(0.0-0.0) | Z=-4.0<sup>b</sup><br>**P=0.000** | 0.0<br>(0.0-0.0) | Z=-2.7<sup>c</sup><br>**P=0.000** |
| Weight loss | 100.0<br>(0.0-100.0) | Z=-7.7<sup>b</sup><br>**P=0.000** | 0.0<br>(0.0-100.0) | Z=-1.7<sup>b</sup><br>P=0.095 | -100.0<br>(-100.0-0.0) | Z=-6.8<sup>c</sup><br>**P=0.000** |
| Weight gain | 0.0<br>(0.0-0.0) | Z=-1.6<sup>b</sup><br>P= 0.102 | 0.0<br>(0.0-0.0) | Z=-1.0<sup>b</sup><br>P=0.317 | 0.0<br>(0.0-0.0) | Z=-.8c<br>P=0.414 |

<sup>a</sup>. Wilcoxon Signed Ranks Test
<sup>b</sup>. Based on negative ranks.
<sup>c</sup>. Based on positive ranks.
P<0.05 are in **Bold** numbers

The changes in HRQOL assessed by EORTC QLQ-C30 are presented in Table 5. The changes for all the parameters from baseline to three months after RT were statistically significant except for the ‘*Nausea and vomiting*’ domain, *‘Dyspnoea’*, ‘*Constipation’* and *‘Diarrhoea’* items. The changes in HRQOL measured by all the domains and items of EORTC QLQ-C30 were found to be significantly different at the three time points except ‘*Diarrhoea’*.

**Table 5:** Comparison of Change of EORTC QLQ-C30 Scores (Change of OHRQOL) of the Sample.

| Domains and items | Baseline to last week of RT course (n=86) |  | Baseline to three months after RT(n=75) |  | Last week of RT course to three months after RT (n=75) |  |
| --- | --- | --- | --- | --- | --- | --- |
|  | Median (IQR) | Sig.<sup>a</sup> | Median (IQR) | Sig.<sup>a</sup> | Median (IQR) | Sig.<sup>a</sup> |
| Global health status | -66.7<br>(-83.3- -50.0) | Z= -7.1<sup>b</sup><br>**P= 0.000** | -16.7<br>(-50.0-0.0) | Z= -5.1<sup>b</sup><br>**P= 0.000** | 33.3<br>(0.0-66.7) | Z= -6.2<sup>c</sup><br>**P= 0.000** |
| Physical functioning | -46.7<br>(-61.7- -40.0) | Z= -7.7<sup>b</sup><br>**P= 0.000** | -26.7<br>(-46.7- -6.7) | Z= -6.0<sup>b</sup><br>**P= 0.000** | 20.0<br>(3.3-40.0) | Z= -4.2<sup>c</sup><br>**P= 0.000** |
| Role functioning | -66.7<br>(-100.0- -50.0) | Z= -7.8<sup>b</sup><br>**P= 0.000** | -50.0<br>(-66.7- -16.7) | Z= -6.5<sup>b</sup><br>**P= 0.000** | 33.3<br>(0.0-50.0) | Z= -4.9<sup>c</sup><br>**P= 0.000** |
| Emotional functioning | -33.3<br>(-66.7-0.0) | Z= -6.6<sup>b</sup><br>**P= 0.000** | 0.0<br>(-50.0-0.0) | Z= -4.1<sup>b</sup><br>**P= 0.000** | 25.0<br>(0.0-50.0) | Z= -2.7<sup>c</sup><br>**P= 0.007** |
| Cognitive functioning | -33.3<br>(-62.5- -16.6) | Z= -7.4<sup>b</sup><br>**P= 0.000** | 0.0<br>(-33.3-0.0) | Z= -4.9<sup>b</sup><br>**P= 0.000** | 16.7<br>(0.0-33.3) | Z= -3.2<sup>c</sup><br>**P= 0.001** |
| Social functioning | -50.0<br>(-66.7- -33.3) | Z= -7.3<sup>b</sup><br>**P= 0.000** | -16.7<br>(-50.0-0.0) | Z= -4.9<sup>b</sup><br>**P= 0.000** | 16.7<br>(0.0-50.0) | Z= -3.9<sup>c</sup><br>**P= 0.000** |
| Fatigue | 66.7<br>(44.4-77.8) | Z= -7.9<sup>c</sup><br>**P= 0.000** | 33.3<br>(11.1-66.7) | Z= -6.4<sup>c</sup><br>**P= 0.000** | -33.3<br>(-44.4-0.0) | Z= -5.3<sup>b</sup><br>**P= 0.000** |
| Nausea and vomiting | 0.0<br>(0.0-0.0) | Z= -3.5<sup>c</sup><br>**P= 0.000** | 0.0<br>(0.0-0.0) | Z= -0.5<sup>c</sup><br>P= 0.595 | 0.0<br>(0.0-0.0) | Z= -2.9<sup>b</sup><br>**P= 0.004** |
| Pain | 66.7<br>(50.0-83.3) | Z= -7.8<sup>c</sup><br>**P= 0.000** | 33.3<br>(16.7-50.0) | Z= -5.9<sup>c</sup><br>**P= 0.000** | -33.3<br>(-50.0-0.0) | Z= -5.3<sup>b</sup><br>**P= 0.000** |
| Dyspnoea | 0.0<br>(0.0-0.0) | Z= -3.1<sup>c</sup><br>**P= 0.002** | 0.0<br>(0.0-0.0) | Z= -1.2<sup>c</sup><br>P= 0.250 | 0.0<br>(0.0-0.0) | Z= -0.7<sup>b</sup><br>P= 0.466 |
| Insomnia | 33.3<br>(0.0-66.7) | Z= -6.8<sup>c</sup><br>**P= 0.000** | 0.0<br>(0.0-33.3) | Z= -3.0<sup>c</sup><br>**P= 0.003** | -33.3<br>(-66.7-0.0) | Z= -5.0<sup>b</sup><br>**P= 0.000** |
| Appetite loss | 66.7<br>(66.7-100.0) | Z= -7.8<sup>c</sup><br>**P= 0.000** | 33.3<br>(0.0-66.7) | Z= -5.9<sup>c</sup><br>**P= 0.000** | -33.3<br>(-66.7-0.0) | Z= -5.4<sup>b</sup><br>**P= 0.000** |
| Constipation | 33.3<br>(33.3-66.7) | Z= -7.3<sup>c</sup><br>**P= 0.000** | 0.0<br>(0.0-33.3) | Z= -1.9<sup>c</sup><br>P= 0.056 | -33.3<br>(-66.7-0.0) | Z= -6.2<sup>b</sup><br>**P= 0.000** |
| Diarrhoea | 0.0<br>(0.0-0.0) | Z= -0.4<sup>b</sup><br>P= 0.705 | 0.0<br>(0.0-0.0) | Z= -1.4<sup>b</sup><br>P= 0.157 | 0.0<br>(0.0-0.0) | Z= -1.0<sup>b</sup><br>P= 0.317 |
| Financial difficulties | 33.3<br>(0.0-66.7) | Z= -6.5<sup>c</sup><br>**P= 0.000** | 0.0<br>(0.0-33.3) | Z= -4.5<sup>c</sup><br>**P= 0.000** | 0.0<br>(-33.3-0.0) | Z= -2.7<sup>b</sup><br>**P= 0.006** |

<sup>a</sup>. Wilcoxon Signed Ranks Test
<sup>b</sup>. Based on positive ranks.
<sup>c</sup>. Based on negative ranks.
P<0.05 are in **Bold** numbers

## Discussion

To our knowledge, this is the first prospective study conducted in oral cancer patients who received RT with or without chemotherapy to report HRQOL in Sri Lanka. The HRQOL was assessed by using EORTC QLQ-H&N35 and EORTC QLQ-C30 questionnaires and the major findings were as follows. Mean scores of all the domains and items in EORTC QLQ-H&N35 and symptom domains and items in EORTC QLQ-C30 were higher during last week of RT compared to the baseline. This is similar for median values except *‘Feeding tube’* and *Weight gain’* items which remained the same. The functional domains of EORTC QLQ-C30 showed the same pattern inversely except for ‘*Role functioning’, these* scores continuously reduced from baseline to three months after RT. The results were suggestive of deterioration of HRQOL within three months period after RT. Almost all the changes of the HRQOL were statistically significant. The results indicate patients suffer from more symptoms after the RT than before RT even though the preference is that the treatment improves HRQOL.^19,20^

A considerable percentage of the sample (17 %) consisted of patients aged more than 70 years. This may be due to the inclusion of the oral cancer patients whose surgical management was not possible due to the old age and therefore, the RT or chemo-RT had been the treatment of choice.^20^ A prospective study conducted among head and neck cancer patients who receive RT had shown slightly different results in the scores of EORTC QLQ-H&N35. ‘*Dry mouth’, ‘Sticky saliva’, ‘Teeth’* and ‘*Opening mouth’* have increased three months after RT compared to baseline and one month after RT whereas in the present study, the corresponding scores three months after RT were lower than the last week of RT. In contrast to the present study the baseline symptom scores were also higher.^21^ That may be due to treatment variation and variation in the site of cancer as the study had been conducted in head and neck cancer patients and two-thirds of the sample had undergone surgery before RT.^21^

Another study reported the mean scores using EORTC QLQ-H&N35 and EORTC QLQ-C30 at baseline, 40 days after initial treatment and one month after RT in much similar manner to the scores of the present study, despite of having done the study in head and neck cancer. In that study, 40 days after initial treatment was similar to last week of RT in the present study as the most patients underwent RT for 30-33 days. The HRQOL of head and neck cancer had reduced from baseline to 40 days. The HRQOL had become better than 40 days compared to one month after RT hence it was more deteriorated than the baseline.^22^

All the functional and symptom domains and items have changed more than 20 points from baseline mean value to last week of RT in EORTC QLQ-C30 which indicates ‘very much change’ in HRQOL and *“Appetite loss”* falls to ‘moderate change’ category according to Osoba et al.^23^ Hence, there is a clinically significant reduction in HRQOL in all the domains and items of EORTC QLQ-C30 according to King, who suggested a change of 10 points to consider as important clinically.^24^ Furthermore, the deterioration of HRQOL from baseline to last week of RT was statistically significant except *“Diarrhoea”* and *“Weight gain”* items.

Of note, the symptoms directly related to the oral cavity were the mostly affected domains and items even after three months of RT. Similarly, *‘Swallowing’, ‘Sticky saliva’, ‘Opening mouth’, ‘Dry mouth’*, and *‘Teeth’*, showed as highly affected parameters in other studies.^22,25^

In contrast to our study findings, another study results revealed that any of the functional domains in EORTC QLQ-C30 had not changed significantly and only the *“Fatigue”, “Pain”, “Insomnia”* and *“Appetite loss”* changed significantly at six weeks compared to baseline. The disparities in the cancer sites, commonest stage and the RT technique used in two samples, may be the reasons.^26^ The present study sample had undergone conventional RT (Cobalt 60) and the majority of studies show a reduction in toxicity and better improvement with time than the immediate effect when RT was given using linear accelerators compared to conventional RT.^27^

The results of this study will be useful to develop treatment guidelines and the areas that healthcare professionals should additionally focus on when managing patients. Patients who receive RT with or without chemotherapy face more difficulties to cope up with the side effects of RT towards the end of the RT course. Both patients and their caregivers need extra support from health care professionals to overcome these problems.^28–30^ There are some limitations of this study. The exact time of assessing baseline HRQOL varied from patient to patient from a few days to just before RT. The tool EORTC QLQ-H&N35 we used, was not specifically designed to assess HRQOL of oral cancer patients. The study was confined only to three months after RT which allows short term evaluation of HRQOL affected by RT with or without chemotherapy. However, high response rate, use of validated commonly used questionnaires, and assess more homogenous group were the strength of this study.

In conclusion, HRQOL of oral cancer patients declined due to RT from baseline to last week of RT and improved three months after RT from last week of RT. Nevertheless, it had not come back to the baseline level from three months after RT. The changes in HRQOL were statistically and clinically significant from baseline to last week of RT, from baseline and three months after RT, and last week to three months after RT.

## Data Availability

All data produced in the present study are available upon reasonable request to the authors

## Prior presentations

Oral presentation and abstract publication at the Annual Scientific Sessions of College of Dentistry and Stomatology of Sri Lanka in 2020 Colombo, Sri Lanka.

## Authors’ disclosures of potential conflict of interest

There is no potential conflict of interest.

## Author contributions

Conception and design: Shamini Kosgallana, Prasanna Jayasekara, Prasad Abeysinghe

Financial support: Shamini Kosgallana

Administrative support: Prasanna Jayasekara, Prasad Abeysinghe

Collection and assembly of data: Shamini Kosgallana

Data analysis and interpretation: Shamini Kosgallana, Prasanna Jayasekara

Manuscript writing: All authors

Final approval of manuscript: All authors

Accountable for all aspects of the work: All authors

## Acknowledgements

We thank all the patients who participated in our study, and their families. We are grateful for all the support provided by the staff of Apeksha Hospital, Maharagama, Sri Lanka. We thank Dr. Nelum Samaruthilaka for advice provided on data analysis.

## Notes

### Competing Interest Statement

The authors have declared no competing interest.

### Funding Statement

This study did not receive any funding

### Author Declarations

Ethical approval to conduct the study was granted from the Ethics Review Committee of Faculty of Medicine, University of Colombo, Sri Lanka (EC-15-200).

